# Sex-stratified phenotyping of comorbidities associated with an inpatient delirium diagnosis using real world data

**DOI:** 10.1101/2023.11.02.23297925

**Authors:** Lay Kodama, Sarah Woldemariam, Alice Tang, Yaqiao Li, Tomiko Oskotsky, Eva Raphael, Marina Sirota

## Abstract

Delirium is a heterogeneous and detrimental mental condition often seen in older, hospitalized patients and is currently hard to predict. In this study, we leverage large-scale, real-world data using the electronic health records (EHR) to identify two cohorts comprised of 7,492 UCSF patients and 19,417 UC health system patients (excluding UCSF patients) with an inpatient delirium diagnosis and the same number of propensity score-matched control patients without delirium. We found significant associations between comorbidities or laboratory test values and an inpatient delirium diagnosis which were validated independently. Most of these associations were those previously-identified as risk factors for delirium, including metabolic abnormalities, mental health diagnoses, and infections. Some of the associations were sex-specific, including those related to dementia subtypes and infections. We further explored the diagnostic associations with anemia and bipolar disorder by conducting longitudinal analyses from the time of first diagnosis of the risk factor to development of delirium demonstrating a significant relationship across time. Finally, we show that an inpatient delirium diagnosis leads to dramatic increases in mortality outcome across both cohorts. These results demonstrate the powerful application of leveraging EHR data to shed insights into prior diagnoses and laboratory test values that could help predict development of inpatient delirium and emphasize the importance of considering patient demographic characteristics including documented sex when making these assessments.

**One Sentence Summary:** Longitudinal analysis of electronic health record data reveals associations between inpatient delirium, comorbidities, and mortality.

## INTRODUCTION

Delirium is a clinical diagnosis defined as fluctuating disturbances in attention, awareness, and cognition that develops over a short period of time and is highly prevalent among older inpatient populations, with an estimated prevalence of 23% in hospitalized older adults (*1, 2*). Both long- and short-term outcomes of delirium are detrimental to patients, distressing to caregivers, and a burden on the healthcare system (*3–5*). Medications for treating delirium are largely for symptomatic management and have not been shown to have clinical benefits and in fact may lead to worse outcomes, especially for older patients (*6*). Prevention is the most effective strategy and often involve non-pharmacological interventions, such as reorientation and cognitive stimulation (*7*), making it imperative to predict which patients may develop delirium using predictive tools to focus prevention efforts for high-risk patients.

Though prevalent, delirium remains challenging to study and predict given its heterogenous clinical nature and diverse risk factors (*2*). Current prediction tools have variable predictive capabilities, with one systematic review finding an area under the receiver operating curve range from 0.52 to 0.94 (*8*). These models have other limitations including validation in small sample sizes or data collected from one institution that may not be generalizable (*9–11*). These models incorporate well-studied risk factors such as existing cognitive impairment and severity of chronic illnesses, but less apparent risk factors also need to be identified to improve these predictive tools (*8, 12, 13*), such as through deep phenotyping of inpatient delirium longitudinally.

Importantly, sex is a major risk modifier for many neurological diseases, with some evidence of sex-differences in delirium (*14–16*). For instance, one study found hypoactive delirium to be more common in female compared to male patients (*15*). Though clinical sex differences have been well-documented in dementia and cognitive impairment -- major risk factors for delirium -- whether sex differences exist in delirium remains largely unstudied due to the lack of sex-stratified studies.

One strategy to overcome these limitations is to employ large-scale, comprehensive real-world data from electronic health records (EHR) combined with robust computational approaches. Data from the EHR have been used to better understand and phenotype complex diseases such as Alzheimer’s disease, type 2 diabetes, and preterm birth (*17–19*). Such deep phenotyping studies can shed insights into clinical risk factors and subtypes of disease as well as point to potential new biological pathways that may be involved in disease.

In this study, we leveraged EHRs from two databases across California to identify differential comorbidities and laboratory test results prior to a patient’s first inpatient admission for delirium, which could serve as potential factors in identifying patients at risk for the condition. Importantly, we also conducted a sex-stratified analysis to identify differences by documented sex in the association between comorbidities or laboratory findings with a delirium diagnosis. We further analyzed specific comorbidities using a longitudinal approach to predict the length of time from the diagnosis of a comorbidity and admission for delirium. Lastly, we conducted a longitudinal analysis to understand the relationship between first inpatient delirium diagnosis and mortality outcomes.

## RESULTS

### Identification of patients with a diagnosis of delirium and their matched controls

We identified 7,492 patients with an inpatient delirium diagnosis and 7,492 propensity-score (PS)-matched control patients within the University of California, San Francisco (UCSF) de-identified electronic health record (EHR) database (∼5 million patients total). Control patients were matched on the following demographic and inpatient visit features: age at admission, patient-identified race, sex, death during admission, inpatient stay length, years of available EHR data, total number of inpatient visits prior to the visit of interest, total number of comorbidities prior to the visit of interest, and whether the visit of interest was in the ICU setting. The visit of interest corresponded to the visit where an inpatient delirium diagnosis was made for the delirium group or a randomly selected inpatient visit for the control group (**Fig. 1**). Similarly, a separate cohort of 19,417 patients with a prior diagnosis of inpatient delirium and 19,417 PS-matched control cohort were identified from the UC-Wide EHR database (∼8.6 million patients total, data from UC Davis, UC Los Angeles, UC Irvine, UC San Diego) with an additional matching criterion that included UC location (**Fig. 1**).

**Fig. 1.**
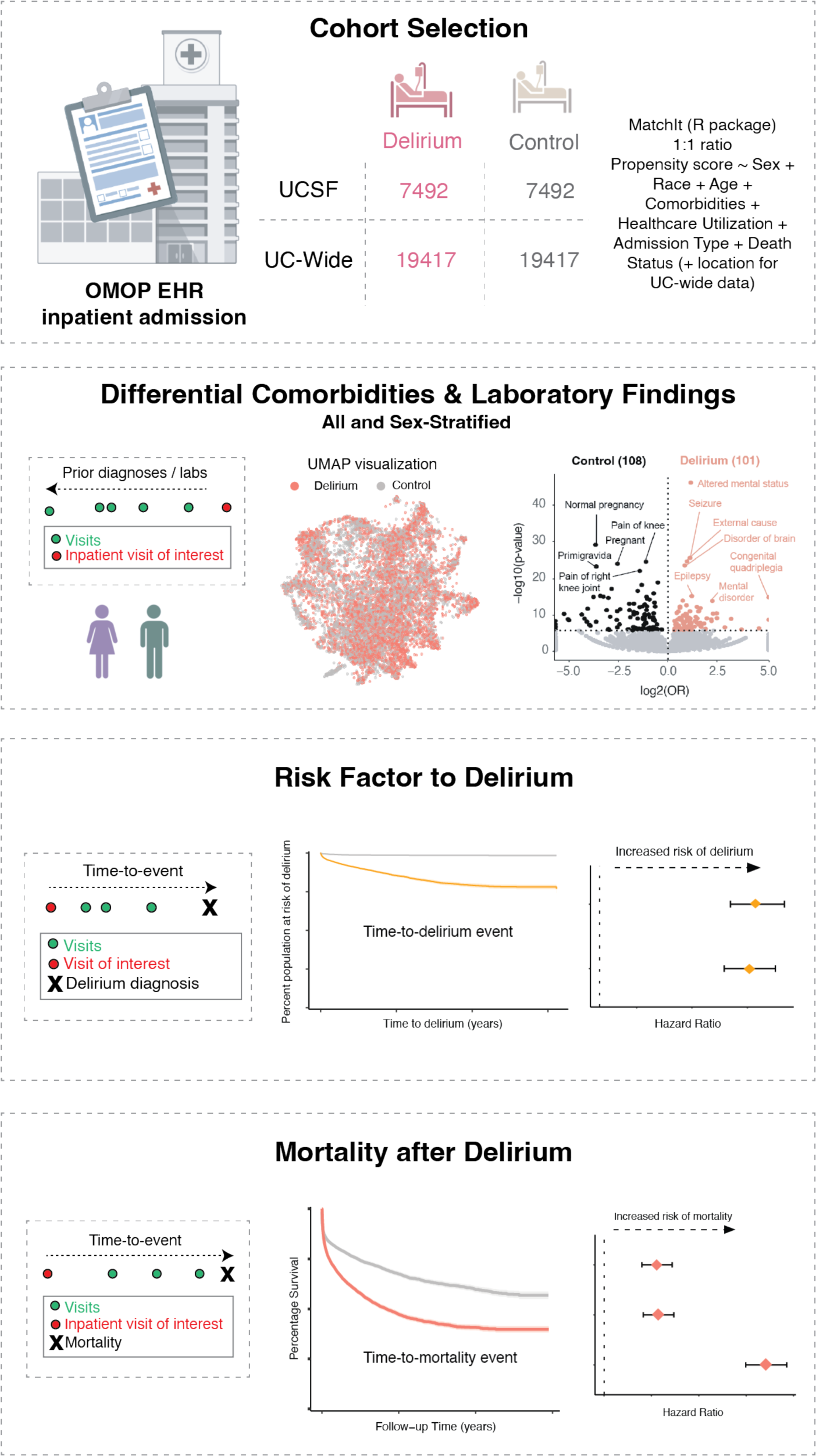
Schematic of study. Patient cohorts were selected from the OMOP electronic health record databases from UC San Francisco (UCSF) and UC-Wide databases (4 sites: UC Davis, UC San Diego, UC Los Angeles, UC Irvine). All patients and their first inpatient diagnosis of delirium were selected and a control cohort with no delirium diagnosis was selected using propensity score matching on the features listed in the formula. Association and longitudinal analyses were done using these cohorts. Association studies were done with prior diagnoses and prior laboratory results, with and without sex- stratification. Longitudinal analyses included time-to-delirium diagnosis after first-time diagnosis of selected potential diagnostic risk factor and time-to-mortality outcome after an inpatient delirium diagnosis.

Post-matching analysis showed adequate matching of covariates with similar PS distributions between the groups (**fig. S1a, c**) and absolute standardized mean differences less than 0.1, except for inpatient stay length which had a wide distribution in both databases (**fig. S1b, d**). Detailed demographic and inpatient visit features for the cohorts generated are shown in Table 1. We defined inpatient delirium broadly using the Observational Medical Outcomes Partnership (OMOP) concept ID 373995 (corresponding to “Delirium”) and excluded diagnoses that had specific causes of delirium in the diagnosis name (such as alcohol-induced delirium or other substance-induced delirium). Using this definition, we were able to capture 84% and 89% of all delirium-related visits in the UCSF and UC-wide databases, respectively, with delirium prevalence within the wide range of published estimates for patients 65 years and older (9.5% mean, 8.9% SD in UCSF data; 3.5% mean, 1.9% SD in UC-wide data) (**fig. S1e, f**).

**Table 1.**
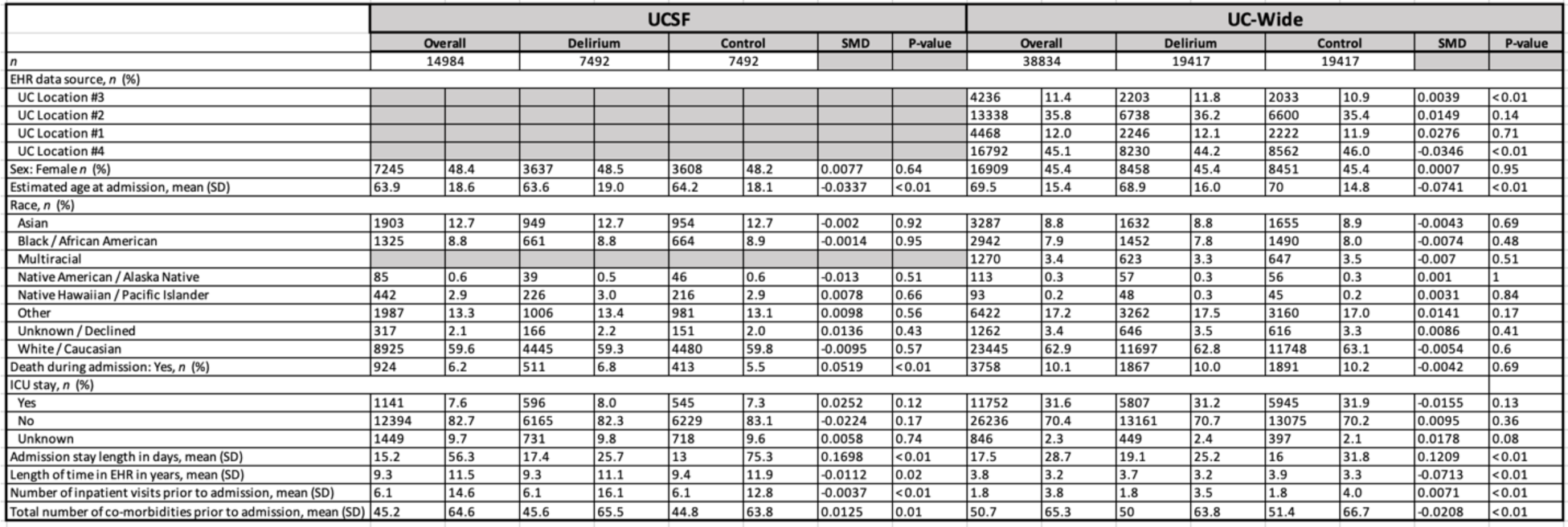
Demographic information of matched cohorts. Table of demographic information for patient cohorts identified in UCSF (left) and UC-wide (right) data. Chi-squared test used for categorical measures. Student’s t-test used for continuous measures. SMD = standardized mean difference.

### Patients with delirium are more likely to be diagnosed with diseases of the nervous system, mental health, metabolic disorders, and infections compared to control patients

To understand potential risk factors associated with an inpatient delirium diagnosis, we first collected all first-time diagnoses made during visits prior to the inpatient admission of interest. Low-dimensional Uniform Manifold Approximation and Projection (UMAP) representation of all non-delirium diagnoses (19,590 features, SNOMED concept IDs) shows a statistically significant separation of patients with a delirium diagnosis versus matched controls by two-sided Mann-Whitney U test (UMAP 1, p-value < 2.2 e-16; UMAP 2 p-value 0.0086; **Fig. 2a**).

**Fig. 2.**
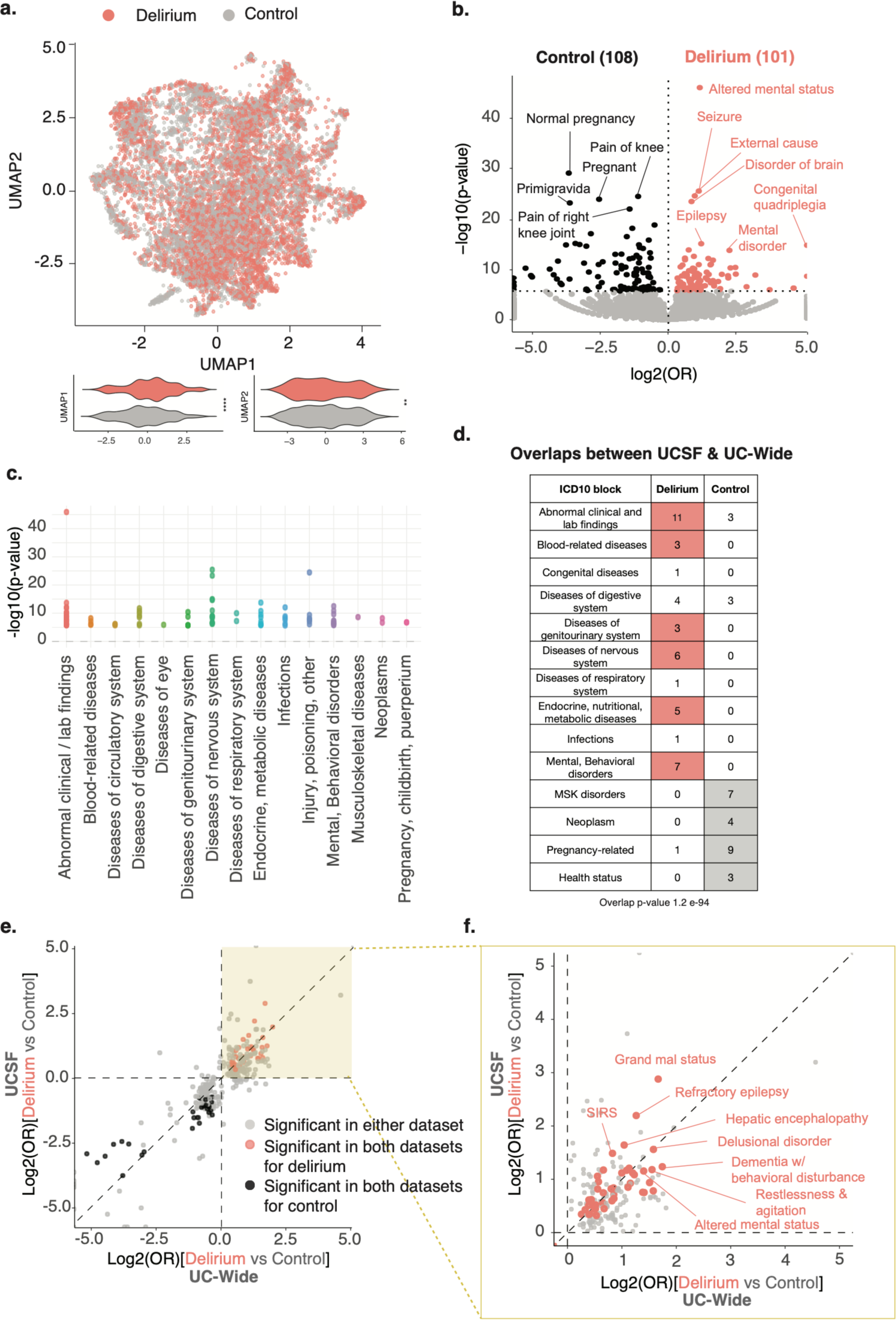
Diagnostic associations with delirium show enrichment of known risk factors of delirium. a. UMAP representation of all first-time, non-delirium diagnoses prior to the inpatient visit of interest. Each dot represents a patient (salmon = patient with delirium, grey = control patient). Violin plots showing distribution of patients across UMAP component 1 (left) and 2 (right). P-values determined by two-sided Mann–Whitney U- tests. **** = p-value < 2.2e-16; ** = p-value 0.0086. b. Volcano plot of differential comorbidities, with diagnoses enriched in controls in black (108 diagnoses) and in delirium patients in salmon (101 diagnoses) and non-significant diagnoses in grey. Significance determined by two-sided Fisher’s exact test with Bonferroni-corrected p- value < 0.05 (at dotted horizontal line). OR = odds ratio. Most significant diagnoses highlighted by name. c. ICD10-diagnostic block representation of significant differential comorbidities identified in (b) for patients with delirium. d. Table showing number of diagnoses overlapping between UCSF and UC-Wide datasets in each ICD10 block for each patient group. Entries with at least 3 or more diagnoses in one patient group compared to the other are colored. Hypergeometric test used to evaluate overlap between the two datasets (p-value 1.2 e-94). e. Log-log plot comparing differential diagnoses between UCSF and UC-Wide databases. Only plotting diagnoses significant in either (grey) or both (salmon) databases. Spearman correlation *ρ* = 0.94 when looking at points significant in both datasets. f. Zoomed in plot of the yellow-highlighted portion of plot in (e). Significant comorbidities found in both databases with diagnoses with the largest odds ratios highlighted.

Differential association analyses of these comorbidities using Fisher’s exact test showed enrichment of distinct comorbidities for patients with delirium compared to control patients, with 101 diagnoses significantly enriched in patients with delirium versus 108 in controls, out of 19,583 diagnoses tested (**Figure 2b**). Control patients had enrichment of diagnoses largely related to pregnancy and other health statuses as well as age-related musculoskeletal diagnoses such as pain in joints and osteoarthritis and skin findings such as melanocytic nevus (**Fig. 2b, c, table S1**). Meanwhile, patients with delirium had enrichment of diagnoses related to diseases of the nervous system, including epilepsy and seizures, mental health and behavioral disorders, and acute diagnoses such as metabolic diseases and infections (**Fig. 2b, c, table S1**). Similar diagnostic associations were seen in the UC-wide cohort based on a hypergeometric test (p-value 1.2 e-94; **Fig. 2d-f, fig. S2**, **table S2, table S3**). Categorizations of these overlapping diagnoses between the two databases by ICD10 diagnostic blocks showed enrichment of diagnoses related to certain disease categories in patients with delirium versus control patients, including blood-related diseases such as anemia, diseases of the genitourinary system such as urinary tract infections, diseases of the nervous system such as epilepsy, metabolic diseases such as hyponatremia, and mental health-related disorders such as bipolar disorder (**fig. S2d, table S3**). These findings are largely consistent with previously-identified risk factors for delirium (*2*).

### Differential laboratory findings corroborate differential associations between comorbidities and delirium

We also conducted enrichment analysis for mean laboratory values and vital signs collected before the inpatient admission of interest. Patients with a delirium diagnosis had significantly higher mean values of certain liver function tests, such as alkaline phosphatase and aspartate transferase, compared to controls in both UCSF and UC-wide datasets, suggesting potential liver dysfunction in these patients (**Fig. 3a**). Elevated mean vital signs included heart rate and respiratory rate **(Fig. 3a**). Meanwhile, glomerular filtration rate and urine creatinine levels were decreased in patients with delirium compared to controls, consistent with kidney dysfunction (**Fig. 3a).** Hemoglobin, hematocrit, and erythrocyte counts were also significantly decreased in patients with delirium compared to controls, consistent with the association with an anemia diagnosis in these patients (**Fig. 3**). The UC-wide dataset also captured certain clinical test scores, including results from the Patient Health Questionnaire (PHQ)-2 and PHQ-9, consistent with the associations with depression in these patients (**table S4**).

**Fig. 3.**
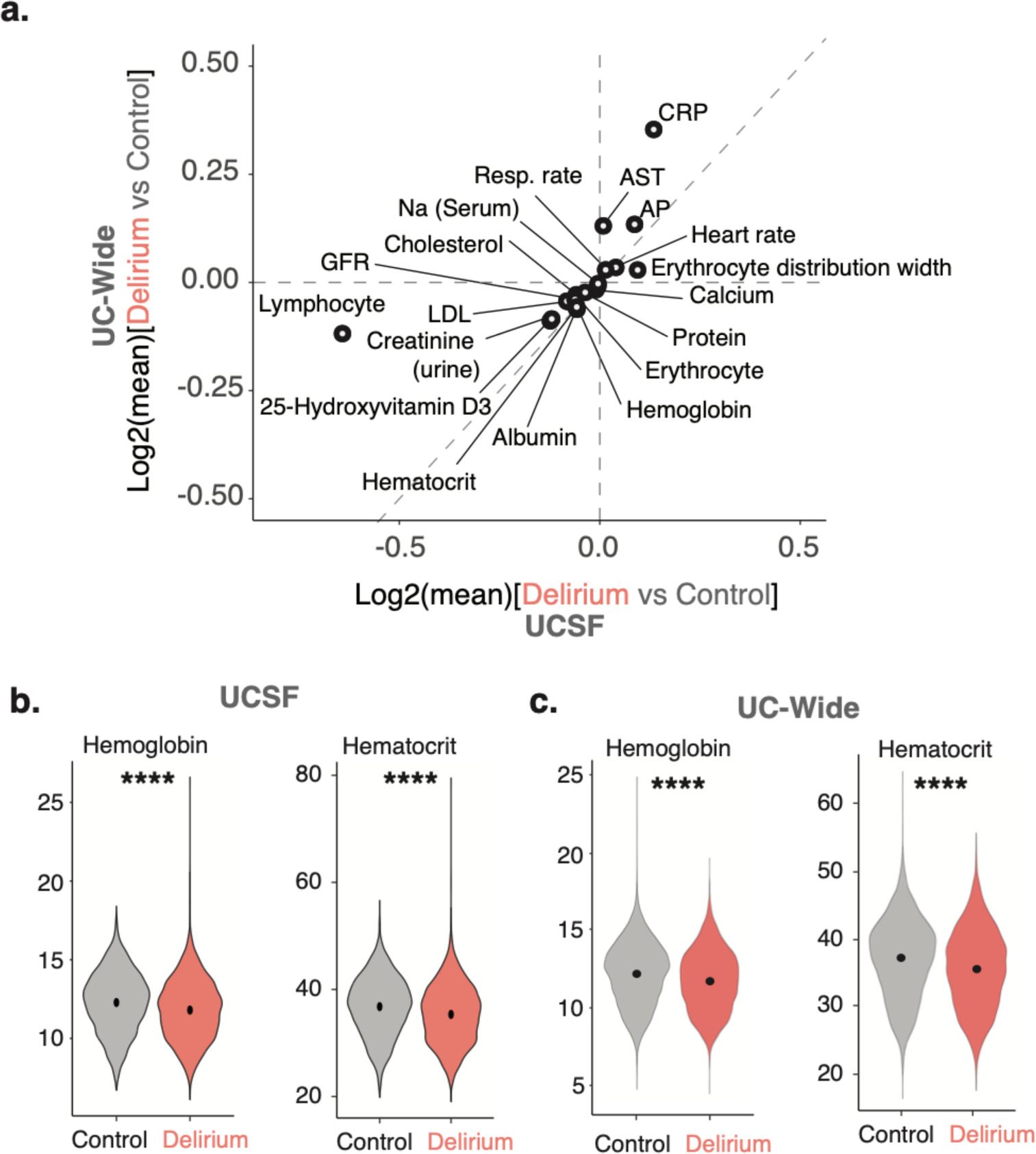
Identification of differential laboratory tests between delirium and control patients. a. Log-log plot comparing all significantly differential laboratory results between delirium versus control patients in UCSF (y-axis) and UC-Wide (x-axis) databases. Axes reflect log base 2 of the mean lab values for delirium versus control patients. b-c. Violin plots showing distribution of patients and their mean hemoglobin and hematocrit values for UCSF (b) and UC-wide (c) datasets. Black point denotes mean of population. Two-sided Mann-Whitney U-test, **** = p-value < 2.2e-16.

### Sex-stratified analysis shows certain infections and dementia subtypes are sex-specific risk factors for delirium

To understand whether any of the comorbidities associated with delirium that we identified are sex-specific, we conducted a sex-stratified association analysis using the same cohort identified above (**fig. S3, fig. S4**). We identified several diagnoses that were significantly associated with delirium in only females, only males, or in both female and male patients with delirium compared to controls in both UCSF and UC-wide datasets (**fig. S3c-e, fig. S4c-e**) as well as sex-specific laboratory results (**fig. S3f, fig. S4f**).

Diagnoses common to both male and female patients with delirium in both datasets were largely similar to the non-stratified analysis, including symptoms of delirium such as altered mental status, restlessness and agitation, and hallucinations, as well as known organic causes of delirium and altered mental status such as seizures, cognitive disorders and other mental disorders (**Fig. 4c**). Interestingly, female and male patients diagnosed with delirium had sex-specific associations with distinct infections and diseases of the nervous system that were statistically significant in both datasets. For instance, female patients with delirium had associations with encapsulated bacterial infections due to Streptococcal bacteria (OR UCSF 2.38; UC-wide 1.61), *Klebsiella pneumoniae* (OR UCSF 2.95, UC-wide 2.14)*, Escherichia coli* (OR UCSF 1.45, UC-wide 1.81), and enterococcal bacteria (OR UCSF 1.53, UC-wide 1.79), while males had associations with *Clostridioides difficile* infections (OR UCSF 3.89, UC-wide 1.55) (**Fig. 4a, b**). Sex-specific associations with subtypes of dementia were also seen, where females had significant associations with Alzheimer’s disease (OR UCSF 2.06, UC-wide 3.55) and vascular dementias with behavioral disturbances (OR UCSF 1.89, UC-wide 3.29) while males had associations with Diffuse Lewy Body disease (OR UCSF 1.51, UC-wide 4.22) and diagnoses related to symptoms of the disease such as visual hallucinations, falls, difficulty walking / muscle weakness, dysphagia, insomnia, and constipation (**Fig. 4a, b, table S5, table S6**). These sex-specific associations could be because these dementia subtypes are known to have sex-differences in their prevalence already (*14*) or could point to sex-specific ways in which delirium manifest in different patient populations.

**Fig. 4.**
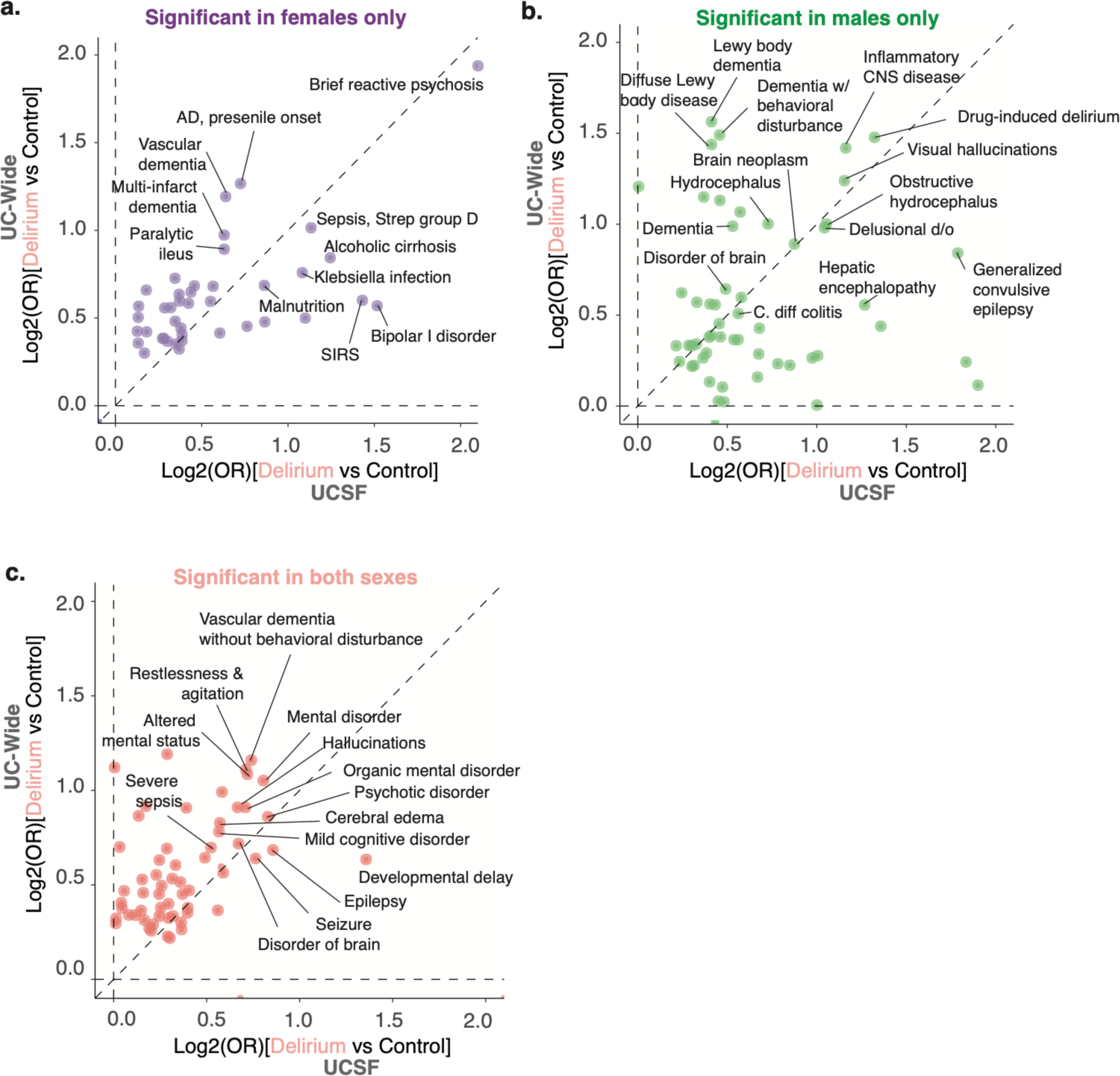
Identification of sex-specific diagnostic associations with delirium, including dementia subtypes and infections. Log-log plots comparing differential diagnostic associations with delirium in UCSF (x-axis) versus UC-wide (y-axis) databases. Plots split by diagnoses significant only in females (a), only in males (b), or in both (c). Diagnoses with largest OR values highlighted. Spearman correlation *ρ* = 0.9 (a), 0.35 (b), 0.55 (c).

Though there were several sex-specific laboratory results found in the UCSF and UC-wide datasets, none were common between the two datasets. Laboratory findings associated with delirium in both male and female patients as well as between both datasets were similar to the results of the non-sex-stratified analysis, including decreased hemoglobin and hematocrit consistent with an anemia diagnosis, decreased GFR consistent with kidney dysfunction, elevated liver function tests such as alkaline phosphatase, and elevation of heart rate (**Fig. 3a, fig. S3f, fig. S4f**).

### Longitudinal analysis from diagnosis of a potential risk factor of delirium to an inpatient delirium diagnosis validates comorbidity association study

To further understand several of the comorbidities associated with an inpatient delirium diagnosis identified above, we carried out a longitudinal time-to-event analysis for anemia and bipolar disorder. We chose these diagnoses for analysis given the higher association of blood-related disorders and mental and behavioral disorders in patients with delirium found through our association study (**Fig. 2d**). Patients with a first-time diagnosis of anemia and no prior diagnosis of delirium and a matched control group with no diagnosis of anemia and no prior diagnosis of delirium were identified. Control patients were matched on the following parameters: age at the visit of interest, patient-identified race, documented sex, years of available EHR data, total number of inpatient visits prior to the visit of interest, and total number of comorbidities prior to the visit of interest. Events were defined as admission for delirium, death, or loss to follow-up.

Kaplan-Meier curve visualization of the data showed stratification by anemia diagnosis status, where those with anemia have increased probability of developing first-time inpatient delirium diagnosis (UCSF 3.4%, UC-wide 1.3% of anemia patients) than those without any anemia diagnosis (UCSF 0.3%, UC-wide 0.3% of control patients) over the course of ∼30 years in the UCSF data and ∼11 years in the UC-wide data (**Fig. 5a, fig. S5a**). Cox proportional hazard ratio analysis unadjusted and adjusted for demographics and visit features revealed a significant increased risk of delirium in those with an anemia diagnosis compared to controls (UCSF HR 9.4; 95% CI, 8.1 to 11; UC-wide HR 4.4; 95% CI, 4.1 to 4.7) (**Fig. 5b**). A similar analysis was done for patients with and without Bipolar I Disorder (BD1) in UCSF patients or bipolar disorder (unspecified type) in UC-wide patients. We found that a diagnosis of BD also increased risk of developing first-time inpatient delirium diagnosis (UCSF 1.9%, UC-wide 0.7% of BD patients) than those without a BD diagnosis (UCSF 0.01%, UC-wide 0.01% of control patients) with a HR of 27 (95% CI, 9.9 to 74.4) for UCSF patients and HR of 7.8 (95% CI, 6.0 to 10.0) for UC-wide patients over the course of ∼20 years in the UCSF data and ∼10 years in the UC-wide data (**Fig. 5c,d, fig. S5b**).

**Fig. 5.**
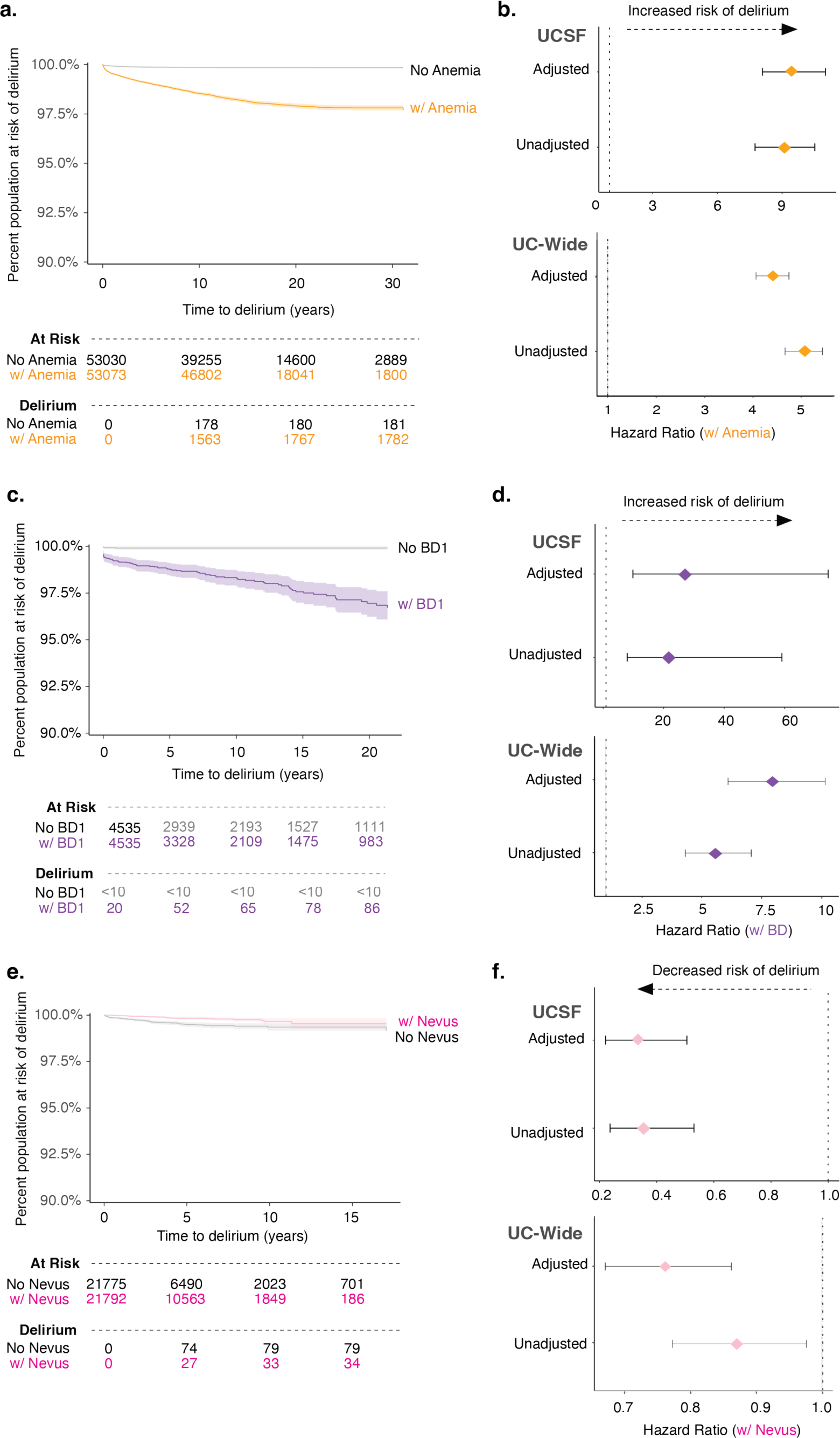
A prior diagnosis of anemia or bipolar disorder leads to increased risk of developing delirium. Kaplan-Meier curve showing time-to-event where event is defined as delirium, death, or loss to follow-up since the first-time diagnosis of anemia (a), bipolar disorder (c), and melanocytic nevus (e) in UCSF patients. Cox proportional hazard ratio analysis done for anemia (b), bipolar disorder (d), and melanocytic nevus (f) with unadjusted and adjusted analyses (adjusted for sex, age at admission, race, length of time in EHR, number of inpatient visits prior, total number of comorbidities, and length of follow-up time in EHR) in UCSF (top) and UC-wide (bottom) patients. Confidence intervals representing 95% CI.

Our association studies also found that certain diagnoses are more enriched in control patients compared to patients with delirium, including diagnoses under the ICD10-CM neoplasm category such as melanocytic nevus (**Fig. 2d, table S3**). We did a similar time-to-event analysis of patients with and without this diagnosis and found that, indeed, a prior diagnosis of melanocytic nevus modestly decreased the risk of developing delirium at a HR of 0.3 (95% CI, 0.2 to 0.5) for UCSF patients and HR of 0.76 (95% CI, 0.67 to 0.86) for UC-wide patients (**Fig. 5e,f, fig. S5c**).

### A single inpatient delirium admission is associated with increased mortality

Previous meta-analyses have documented increased risk of mortality after an inpatient delirium admission (*3*). To validate these findings in a larger population size while also accounting for more covariates than previously tested, including health status, we used our cohort of patients with delirium and their matched controls to conduct a longitudinal time-to-event analysis, where an event was defined as mortality or loss to follow-up. Kaplan-Meier survival curve visualization of the data showed different probabilities of survival rates between patients with an inpatient delirium admission (median 8.47 years; 95% CI, 7.8 to 9.35 in delirium group) versus control patients (**Fig. 6a**). Cox proportional hazard ratio analysis unadjusted and adjusted for demographic characteristics (sex, age at admission, race, length of time in EHR, number of inpatient visits prior, total number of comorbidities) and visit features (type of visit, visit length, and length of follow-up time in EHR) revealed a significant increased risk of delirium in those with a delirium diagnosis compared to controls (HR 1.2; 95% CI, 1.16 to 1.29) (**Fig. 6b**). A similar analysis for the UC-wide patient cohort also showed increased mortality in those with a delirium diagnosis compared to controls with a HR of 1.14 (95% CI, 1.1 to 1.18) (median 8.96 years; 95% CI, 8.16 to 10.1 in control group; median 3.83 years; 95% CI, 3.71 to 4.1 in delirium group) (**Fig. 6c,d**).

**Fig. 6.**
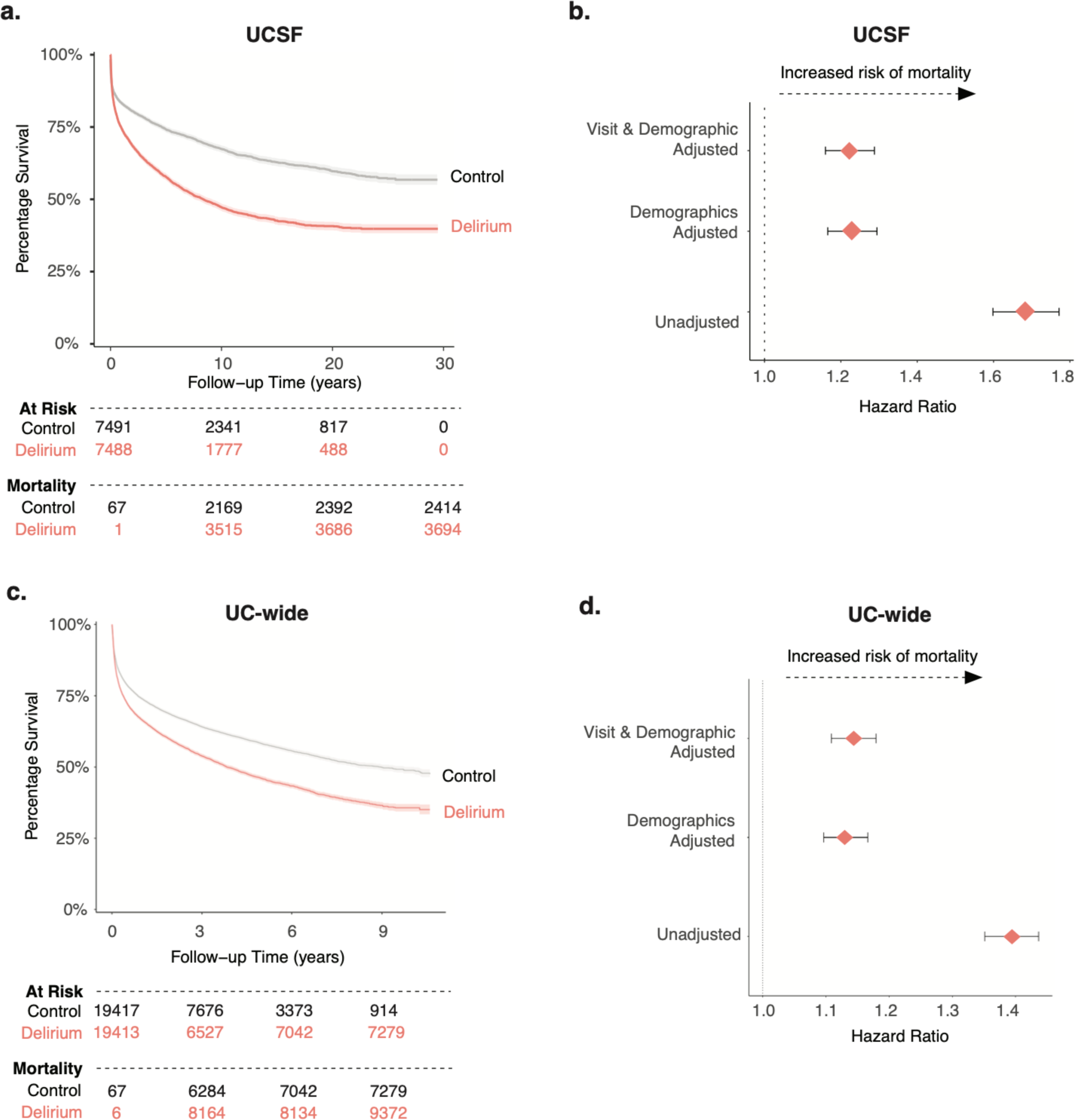
Increased mortality outcome after an inpatient delirium diagnosis. Kaplan-Meier survival curve showing time-to-death after inpatient admission of interest for control (grey) and delirium (salmon) patients in UCSF (a) and UC-Wide (c) data. Follow up time periods differ between the two datasets given the discrepancy in the number of years captured by each dataset. Cox proportional hazard ratio analysis done for in UCSF (b) and UC-wide (d) cohort with unadjusted and adjusted analyses (adjusted for sex, age at admission, race, length of time in EHR, number of inpatient visits prior, total number of comorbidities, type of visit, visit length, and length of follow-up time in EHR).

## DISCUSSION

In the past few decades, clinical data from the EHR combined with integrative computational approaches have enabled the dissection and deep phenotyping of complex, heterogeneous diseases in an unbiased manner. This study aims to apply this strategy to better understand both risk factors and outcomes of patients who are diagnosed with delirium while in the hospital. We found that many well-known risk factors of delirium emerged through our analysis looking at comorbidities associated with an inpatient delirium diagnosis, including chronic neurological conditions such as dementia and epilepsy as well as acute conditions such as infections and metabolic disturbances. Our sex-stratified association studies revealed that, even within these large categories of diseases of the nervous system and infections, subtypes of conditions were sex-specific. We also validated several of these comorbidities associated with delirium, including anemia, bipolar disorder, and melanocytic nevus, through time-to-event analyses. Finally, we showed that a delirium diagnosis is independently associated with an increased risk of mortality, with UCSF and UC-wide data showing a median survival of ∼8.5 and ∼4 years, respectively.

To our knowledge, this is the first study to conduct a deep phenotyping of delirium versus control patients using the EHR data with over 7,000 patients with delirium in the exploratory dataset and ∼20,000 patients in the validation dataset. We aimed to find an appropriately matched control cohort by using propensity score matching and including covariates that assess patient health status as well as frequency of healthcare utilization. We also did not exclude patients based on prior cutoffs such as age thresholds or ICU versus non-ICU admissions, as done by previous studies. This strategy enabled us to include as many delirium cases as possible in order to find potential associations that may be subtle but meaningful. For instance, anemia before admission for delirium was enriched in delirium patients compared to controls in both UCSF and UC-wide datasets. This was also confirmed at the laboratory level, with significant mean hemoglobin difference in patients with and without delirium (11.6 versus 12.2 respectively). Our longitudinal analysis validated our comorbidity analysis and provided an understanding of the time between diagnosis of a risk factor to the development of delirium. In the UCSF data, for instance, roughly 3% of patients with anemia went on to develop delirium within 10 years of the diagnosis while less than 0.5% of patients without anemia developed delirium in this timeframe (hazard ratio of 9.4). One study in 700 patients specifically undergoing lumbar spinal fusion found that perioperative anemia is a risk factor for developing delirium post-operation (*20*). Our study is the first to show that a diagnosis of anemia may be a more generalizable risk factor for developing inpatient delirium. Further studies need to be done to understand whether treating anemia more aggressively will reduce development of delirium.

The large number of patients also enabled us to dissect the diagnostic associations with delirium in a sex-stratified manner. Our findings point to potentially new biological insights into the pathophysiology of delirium that are sex dependent. For instance, male patients had a significant association with *Clostridioides difficile* infections, an infection that often develops after antibiotic use. This could point to either the *Clostridioides difficile* toxin and the downstream consequences of the infection being associated with delirium or the antibiotic that was used that led to the infection being associated with delirium. Meanwhile, *Klebsiella* and *E. coli* infection were enriched in female patients with delirium, organisms that most commonly cause urinary tract infections, a source of infection more common in females compared to males. Dementia, one of the known risk factors of delirium, is also known to have sex-biased features, including in prevalence, clinical progression, and neuropathological findings (*14*). Interestingly, our association studies found that subtypes of dementia were associated with delirium in a sex-dependent manner. Further studies will need to be done to understand whether this is due to the underlying sex differences in the prevalence of these dementia subtypes or sex differences in the pathophysiology of these dementias contributing to delirium.

There are several limitations to our study that need further investigation. Given that illness-severity is a well-studied risk factor for delirium, we used several indirect metrics of health status, including number of inpatient admissions and number of comorbidities, to find an appropriately matched control cohort to our delirium cohort. In this way, we were able to compare groups with similar health statuses in order to extract more subtle risk factors that may be associated with delirium. These metrics of health status are still indirect and could be improved. For instance, though we matched on the number of comorbidities to identify a control cohort with a similar health status, some comorbidities are more functionally debilitating and severe than others. Given our control group had more pregnancy-related diagnoses, this suggests that though the control group may have a similar frequency of healthcare utilization, they are overall healthier than the matched delirium cohort. Future studies could use other health status metrics, such as the Charlson Comorbidity Index or the American Society of Anesthesiologists (ASA) physical status classification score, which classifies patients from 1 to 5, with higher scores denoting patients with severe illness (*21*). ASA was available in our UC-wide dataset and was significantly higher in patients with delirium compared to control patients (**table S4**). The mean values, however, were 2.95 in control patients versus 3.05 in delirium patients, suggesting this difference is likely not clinically meaningful, given the scores are integer numbers.

Defining an accurate delirium diagnosis in the EHR is challenging. Detection and clinical diagnosis of delirium itself is often challenging and likely underdiagnosed (*2, 22*). As a result, the published prevalence of delirium in older patients varies widely, with some estimates around 23% (*23*) while others as high as 88% in palliative care patients (*24*). Prevalence of delirium in our study was lower than these estimates, likely due to our restricted definition of delirium only including one OMOP concept ID and including younger patients. This definition, however, likely missed patients that did not fit this exact delirium definition. Documentation of delirium is unfortunately inconsistent in the EHR even with a confirmed diagnosis, with one study finding that only 9 out of 25 cases coded by ICD-9 code (*25*). Other measures of delirium could be used in future studies to incorporate more patients, such as searching for delirium symptoms in clinical notes or including patients who have been screened for delirium using clinical tools such as the 4 A’s Test (4AT) or the confusion assessment method (CAM) (*26, 27*).

In this study, the definition of sex in the EHR is likely a combination of sex assigned at birth, legal sex, and sex determined by the clinician. Documentation of gender identity remains a challenge in the EHR and only recently has garnered attention to provide more inclusive and affirmative health care for all patients (*28, 29*). Further studies will need to be done to understand whether the associations found in this study are different when taking into context biological sex versus patient-reported gender identity. Similarly, other social determinants of health were not taken into account in this study, such as the patient’s level of formal education, which can influence cognitive reserve, as well as family / caregiver dynamics.

Overall, our study demonstrates the powerful application of the EHR to study heterogeneous disease processes and to better understand the risk factors and outcomes of disease at both a large patient population scale and a longitudinal time course. These results not only confirmed some of the known risk factors of delirium but also generated several new clinical hypotheses that will need to be further investigated. These findings could also help develop future modeling studies for predicting which patients will develop delirium to focus prevention efforts towards these patients. Understanding the impact of sex differences in delirium remains understudied, and this study points to the importance of doing sex-stratified analyses and the potentially interesting pathophysiology of delirium that interacts with sex.

## MATERIALS AND METHODS

### Cohort identification

All analysis of UCSF and UC-wide EHR data was performed under the approval of the Institutional Review Boards. All clinical data were de-identified and written informed consent was waived by the institutions. Patient cohorts were identified using the UCSF de-identified and UC-wide HIPAA-compliant limited data set OMOP EHR databases. The UCSF dataset included over five million patients from January 1, 1982 to February 20, 2023 while the UC-wide dataset included over seven million patients from January 1, 2012 to April 19, 2023 from 4 sites (UC San Diego, UC Los Angeles, UC Irvine, and UC Davis). Patients with inpatient delirium were identified using the OMOP concept ID 373995, SNOMED code 2776000 (corresponding to “Delirium”), filtered for first-time diagnosis of delirium during an inpatient stay (i.e., ‘visit of interest’). The inpatient control cohort with no delirium diagnosis was identified through propensity score (PS) matching (matchit R(*30*)) by a generalized linear model at a 1:1 ratio using a nearest neighbor method and the following matching criteria: assigned sex, patient-reported race, estimated age at admission, years in EHR prior to visit, total number of comorbidities and inpatient visits prior to the visit of interest, stay length, stay type (ICU vs non-ICU), death during admission, and UC location (for UC-wide dataset).

### Differential comorbidity analysis

Comorbidities were identified prior to the visit of interest with the earliest entry of every diagnosis. All patients and their comorbidities were visualized using Uniform Manifold Approximation and Projection (UMAP) using the R-implementation of the umap-learn package from Python (*31*). Correlations between variables (delirium status or sex) and UMAP coordinates were analyzed using Mann-Whitney *U*-tests. Differential comorbidity analysis between patients with delirium and controls was done using Fisher’s exact test and significance determined by Bonferroni-corrected threshold of p-value < 0.05. ICD10-CM blocks were also used to visualize differential comorbidity results. A dictionary was used to match the SNOMED codes to ICD10 codes then mapped to categorical modules using the ICD10 codes. Overlaps between results of analyses using UCSF versus UC-wide datasets were done using hypergeometric test or Spearman correlation. A similar analysis was done for the sex-stratified analysis.

### Differential laboratory test results analysis

Laboratory test results were collected for tests done prior to the visit of interest, and the median values for all numerical lab tests was calculated. Lab tests that had more than 95% of patients missing results were excluded from the analysis. Lab value distributions were compared using Mann-Whitney *U*-tests across delirium status or sex.

### Longitudinal analyses

Time-to-event analysis was done by first identifying patients with a diagnosis of bipolar disorder (BD) and no prior delirium diagnosis. PS-matched control cohorts with no diagnosis of BD were identified using similar matching-criteria as described above. Time-to-event was calculated as time from first-time diagnosis of BD (or another non-BD diagnosis for the control patients) to either first inpatient delirium diagnosis, death, or loss to follow-up. Cox regression model was used to determine the hazard ratio, confidence intervals and significance (survival R(*32*)). Time-to-event analysis for mortality after delirium was done using a similar strategy.

## List of Supplementary Materials

Fig. S1 (Related to Fig. 1): Matching results of delirium and control cohort from UCSF and UC-Wide EHR datasets

Fig. S2 (Related to Fig. 2): Diagnostic associations with delirium in UC-wide data

Fig. S3 (Related to Fig. 4): Diagnostic and laboratory associations with delirium by sex in UCSF data

Fig. S4 (Related to Fig. 4): Diagnostic and laboratory associations with delirium by sex in UC-wide data

Fig. S5 (Related to Fig. 5): Time from diagnostic risk factor to delirium in UC-wide data

Table S1 (Related to Fig. 2): Differential comorbidities between delirium and control patients in UCSF data

Table S2 (Related to Fig. 2): Differential comorbidities between delirium and control patients in UC-wide data

Table S3 (Related to Fig. 2): Differential comorbidities between delirium and control patients significant in both datasets

Table S4 (Related to Fig. 4): Sex-stratified differential comorbidities between delirium and control patients significant in UCSF data

Table S5 (Related to Fig. 4): Sex-stratified differential comorbidities between delirium and control patients significant in UC-wide data

## Acknowledgments

We thank the UCSF Information Commons for their help navigating the UCSF EHR database, especially Evan Phelps, and the UC Health Data Warehouse team for their help with the UC-wide database, especially Robert Follett.

## Author contributions

Conceptualization: L.K., M.S.

Methodology: L.K., S.W., A.T., Y.L., T.O., M.S.

Investigation: L.K., M.S. Visualization: L.K.

Funding acquisition: L.K., M.S. Project administration: L.K., M.S. Supervision: M.S.

Writing – original draft: L.K., M.S.

Writing – review & editing: L.K., S.W., A.T., Y.L., T.O., E.R., M.S.

## Data and materials availability

The UCSF EHR database is available to individuals affiliated with UCSF who can contact the UCSF’s Clinical and Translational Science Institute (CTSI) or the UCSF’s Information Commons team for more information. The UC-wide EHR database is only available to UC researchers who have completed analyses in their respective UC first and have provided justification for scaling their analyses across UC health centers (more details at https://www.ucop.edu/uc-health/departments/center-for-data-driven-insights-and-innovations-cdi2.html or by contacting. The code used for analysis will be made available on github.

---

**Supplementary Figure 1.**
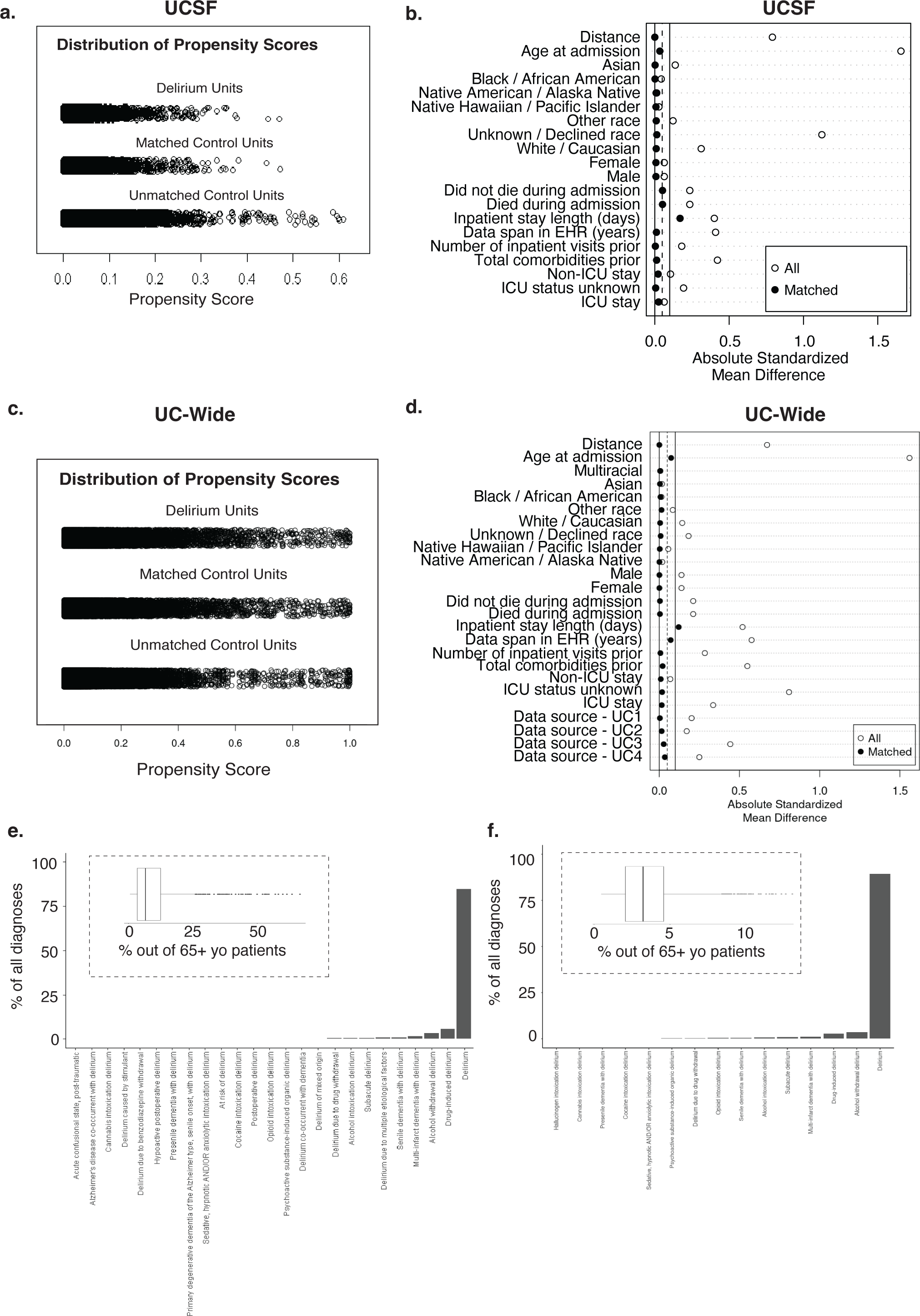
(Related to **Figure 1) Matching results of delirium and control cohort from UCSF and UC-Wide EHR datasets** a-d. Distribution of propensity scores for patients with delirium, matched controls, and unmatched controls for UCSF (a) and UC-wide data (c). Absolute standardized mean difference between matched patients (black dot) versus all patients from the selection pool (white dot) for each demographic parameter used in the matching for UCSF (b) and UC-wide data (d). e-f. Bar plot of the percent of diagnoses out of all delirium-related diagnoses in UCSF (e) and UC-wide data (f). Inset boxplot showing percent of patients with a delirium diagnosis out of all patients admitted to the hospital on a given visit date who are 65 years or older at UCSF (e) and UC-wide (f).

**Supplementary Figure 2.**
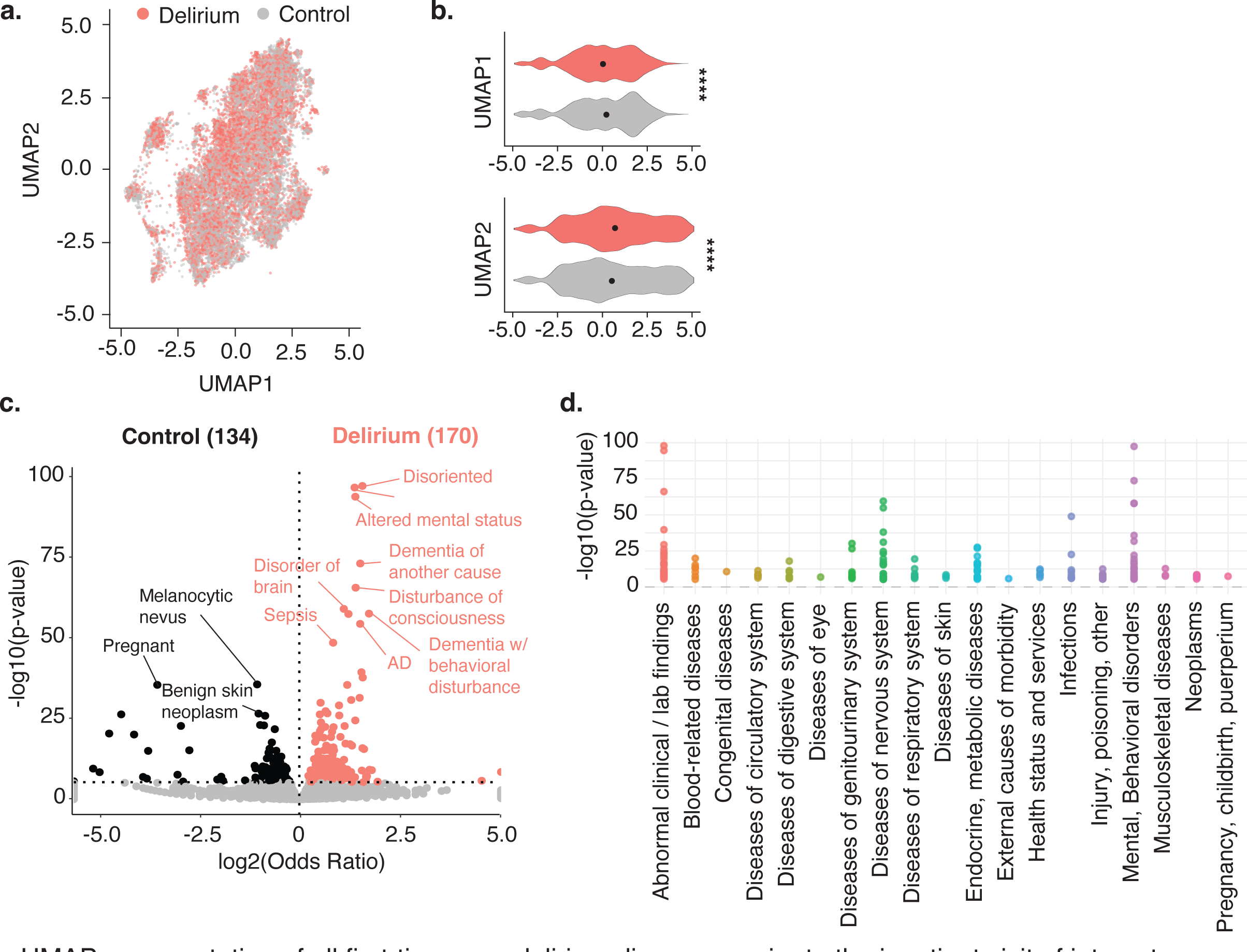
(related to Figure 2): Diagnostic associations with delirium in UC-wide data a.UMAP representation of all first-time, non-delirium diagnoses prior to the inpatient visit of interest. Each dot represents a patient (salmon = patient with delirium, grey = control patient). b. Violin plots showing distribution of patients across UMAP component 1 (top) and 2 (bottom). p-values determined by two-sided Mann–Whitney U-test. **** = p-value 1.5e-12; **** = p-value < 2.2e-16. c. Volcano plot of differential comorbidities, with diagnoses enriched in controls in black (134 diagnoses) and in delirium patients in salmon (170 diagnoses) and non-significant diagnoses in grey. Significance determined by two-sided Fisher’s exact test with Bonferroni-corrected p-value < 0.05 (at dotted horizontal line). OR = odds ratio. Most significant diagnoses highlighted by name. d. ICD10-diagnostic block representation of significant differential comorbidities identified in (c) for patients with delirium.

**Supplemental Figure 3.**
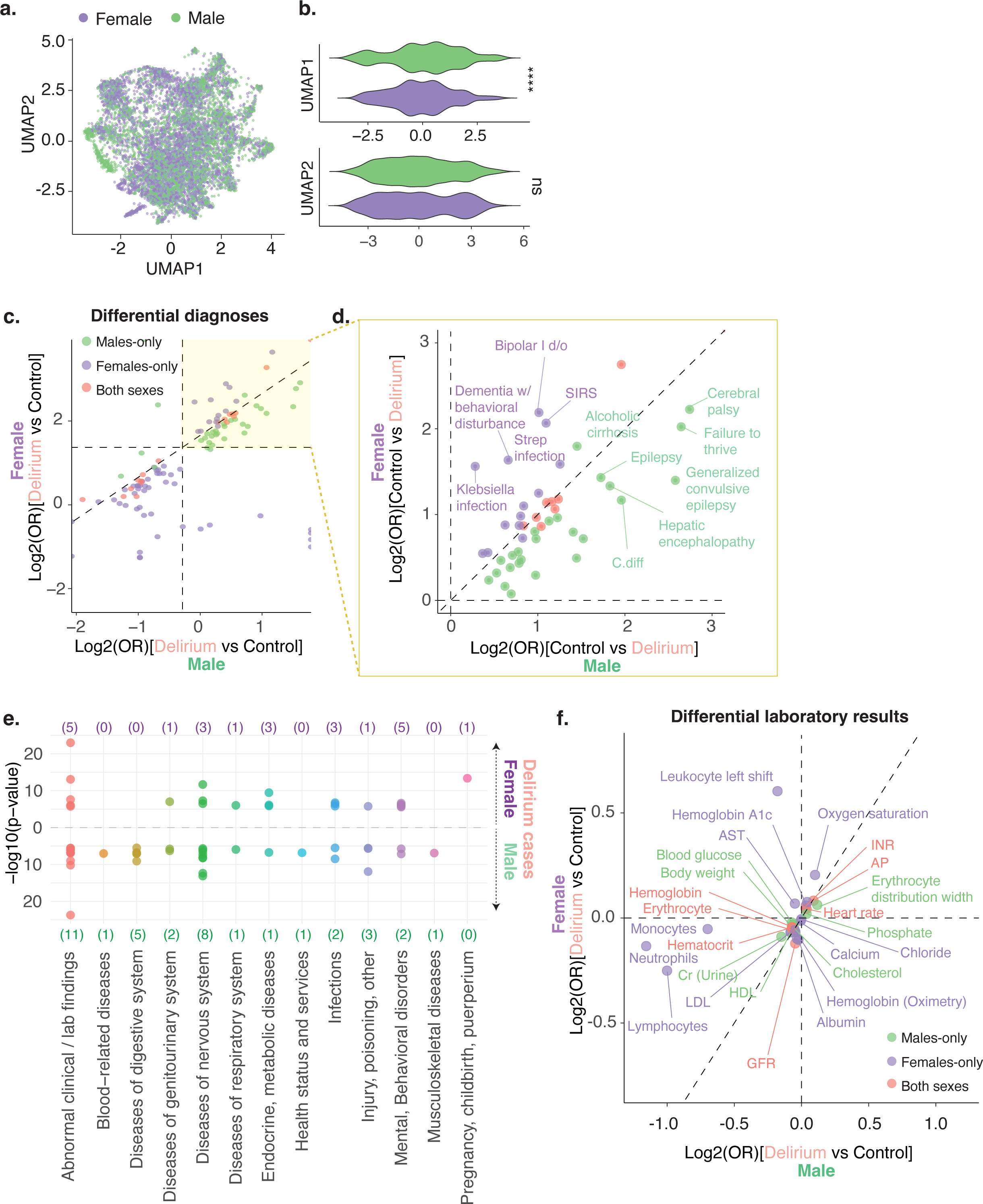
(related to. **Fig 4): Diagnostic and laboratory associations with delirium by sex in UCSF data** a.UMAP representation of all first-time, non-delirium diagnoses prior to the inpatient visit of interest. Each dot represents a patient (purple = female, green = male). b. Violin plots showing distribution of patients across UMAP principle component 1 (top) and 2 (bottom). p-values determined by two-sided Mann–Whitney U-test. **** = p-value < 2.2e-16. c. Log-log plot comparing differential diagnoses between female and male patients. Diagnoses significant in females only (purple), males only (green), or in both sexes (pink). d. Zoomed in plot of the yellow-highlighted portion of plot in (c). Significant comorbidities found in either male or female patients highlighted. e. ICD10-diagnostic block representation of significant differential comorbidities identified in (c) for patients with delirium, female (top), male (bottom). f. Log-log plot comparing differential laboratory results between female and male patients. Laboratory tests significant in females only (purple), males only (green), or in both sexes (pink).

**Supplemental Figure 4.**
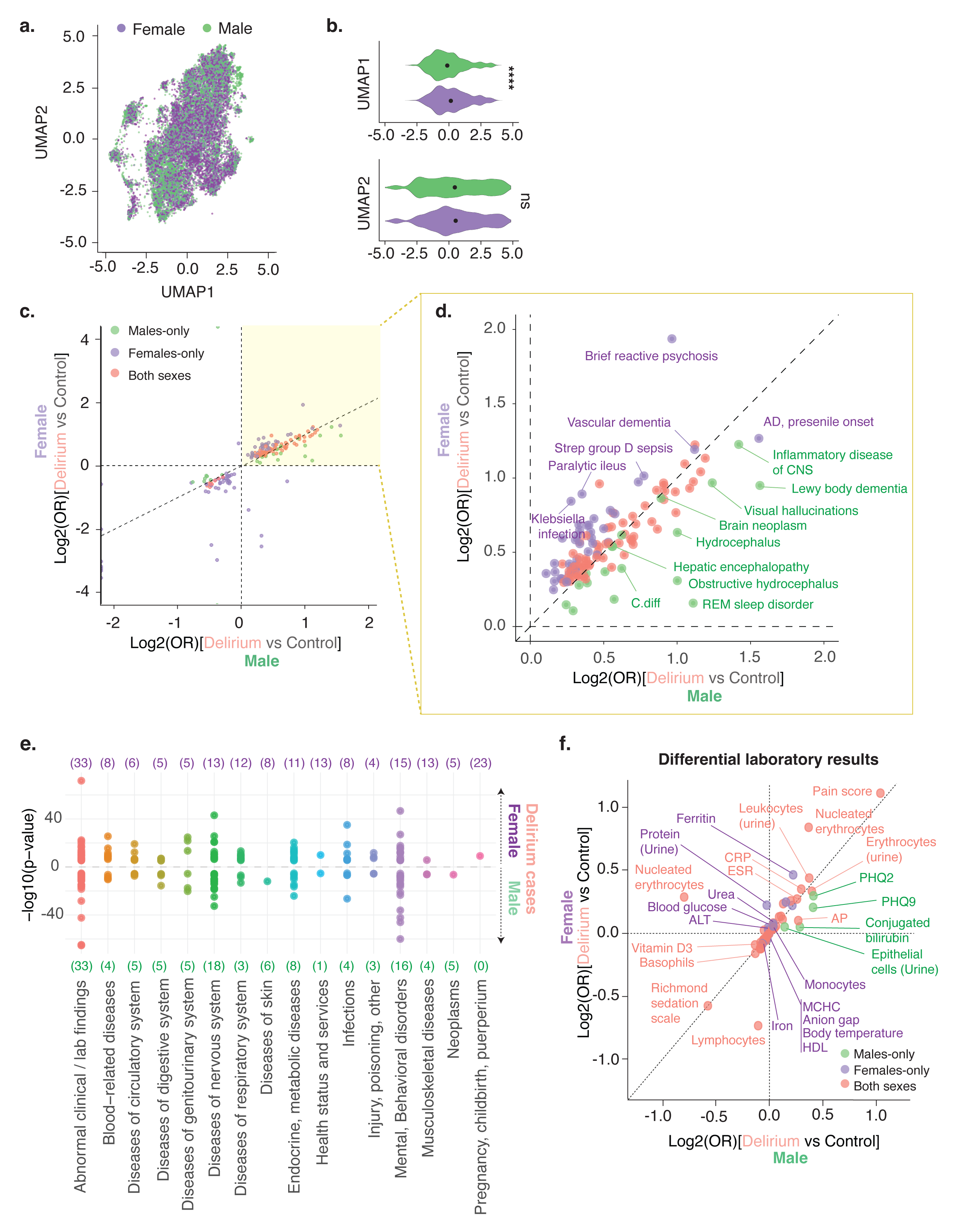
(related to Fig 4): **Diagnostic and laboratory associations with delirium by sex in UC-wide data** a.UMAP representation of all first-time, non-delirium diagnoses prior to the inpatient visit of interest. Each dot represents a patient (purple = female, green = male). b. Violin plots showing distribution of patients across UMAP principle component 1 (top) and 2 (bottom). p-values determined by two-sided Mann–Whitney U-test. **** = p-value = 5.3e-15. c. Log-log plot comparing differential diagnoses between female and male patients. Diagnoses significant in females only (purple), males only (green), or in both sexes (pink). d. Zoomed in plot of the yellow-highlighted portion of plot in (c). Significant comorbidities found in either male or female patients highlighted. e. ICD10-diagnostic block representation of significant differential comorbidities identified in (c) for patients with delirium, female (top), male (bottom). f. Log-log plot comparing differential laboratory results between female and male patients. Laboratory tests significant in females only (purple), males only (green), or in both sexes (pink).

**Supplementary Figure 5.**
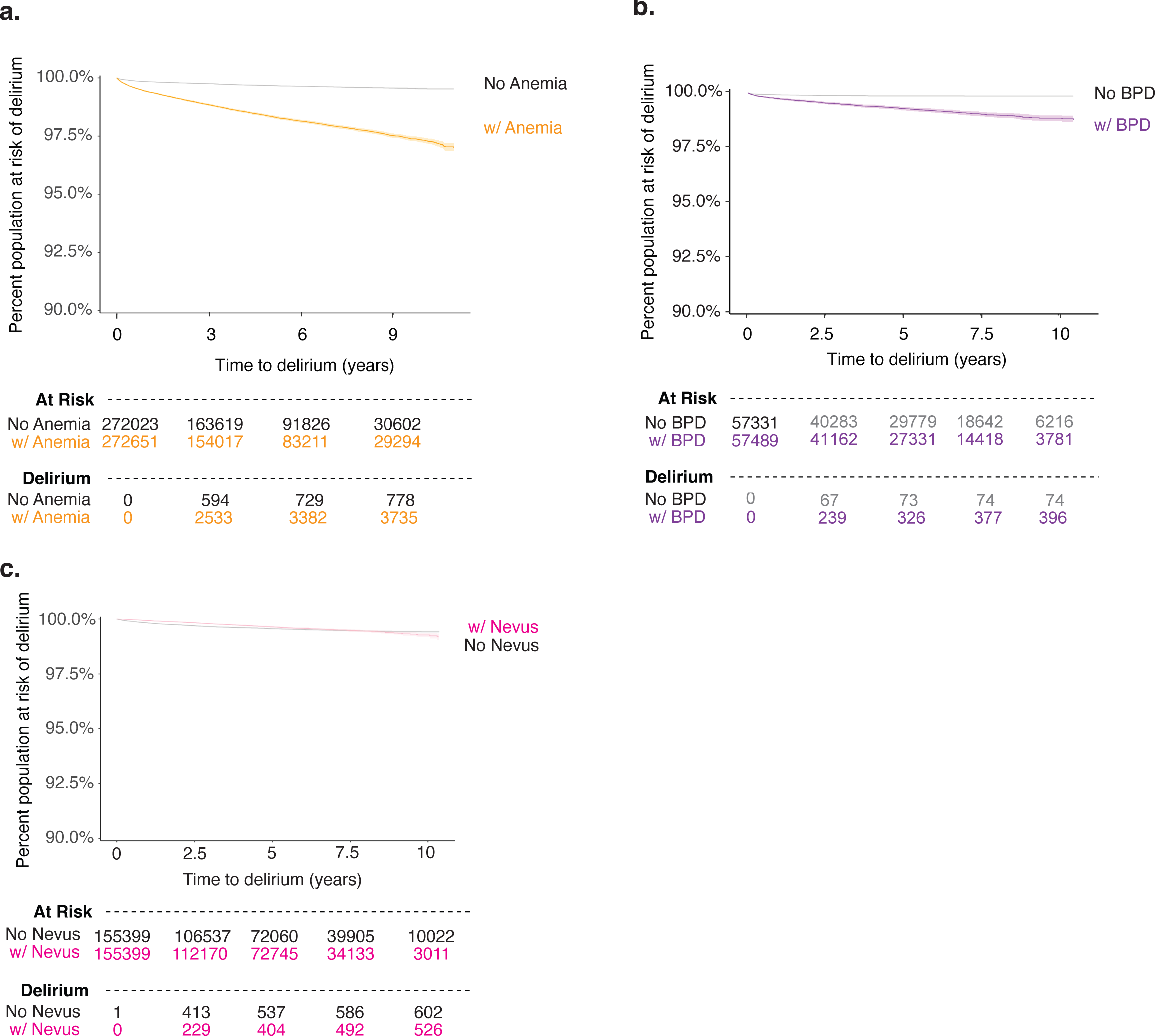
(related to Fig 5). Time from diagnostic risk factor to delirium in UC-wide data Kaplan-Meier curve showing time-to-event where event is defined as delirium, death, or loss to follow-up since the first-time diagnosis of anemia (a), bipolar disorder (b), and melanocytic nevus (c) in UC-wide patients.

